# Potential effect of replacing sedentary behavior with physical activity on the risk of atrial fibrillation: results from the UK Biobank and REGARDS study

**DOI:** 10.64898/2026.10.02.26364589

**Authors:** Linzi Li, Amit J. Shah, Yi-An Ko, Dayna A. Johnson, Yan V. Sun, Chang Liu, Emily B. Levitan, Virginia J. Howard, Suzanne E. Judd, Elsayed Z. Soliman, Alvaro Alonso

**Author notes:** Corresponding author: Linzi Li, Address: Department of Epidemiology, Rollins School of Public Health Emory University 1518 Clifton Rd NE, Atlanta, GA 30322.

## Abstract

**Background:** Sedentary behavior (SB) has been associated with cardiovascular disease risk, independent of physical activity (PA) and sleep. However, the effect of replacing SB with PA or sleep on atrial fibrillation (AF) risk remains unclear.

**Methods:** We studied 86,096 participants from the United Kingdom Biobank (UKB) and 5,777 from the United States (US) REasons for Geographic And Racial Differences in Stroke (REGARDS) prospective cohort studies who were free of AF at baseline and had valid accelerometry data. The two cohorts were included to evaluate the consistency of associations across distinct populations in the UK and the US, two regions with a high burden of AF. Daily time spent in light PA (LPA), moderate to vigorous PA (MVPA), and SB were derived from accelerometer recordings; sleep was accelerometer-derived in UKB and self-reported in REGARDS. Incident AF was identified using ICD-10 codes (I48.x) listed as either a primary diagnosis, secondary diagnosis, or cause of death through hospital and death registry data linkage in UKB and in a follow-up visit through electrocardiogram and self-reported medical history in REGARDS. Isotemporal substitution models were used to examine replacing 30 minutes of SB with LPA or MVPA in both cohorts and with sleep in UKB on the outcome of incident AF.

**Results:** Among the UKB participants [57% female, mean age 56.0 (SD 7.8) years, 98% White race], 4,035 AF cases were identified during a median of 7.9 years of follow-up. Replacing 30 minutes of SB with PA was associated with a slightly lower risk of AF in the minimally adjusted models but the associations were attenuated after full adjustment (LPA: HR 0.99, 95% CI 0.95-1.02; MVPA: HR 1.00, 95% CI 0.99-1.02). The REGARDS participants had an average age of 63 (SD 8.3) years, 55% female, 31% Black and 69% White adults, with 486 AF cases identified. Fully adjusted substitution of 30 minutes of SB with MVPA was associated with lower AF odds (OR 0.80, 95% CI 0.63-1.01), whereas LPA showed no association. In UKB, replacing SB with sleep was not associated with AF risk. The magnitude of the association of MVPA substitution with AF appeared stronger among REGARDS participants aged ≥65 years and those reporting <7 hours/day of sleep.

**Conclusion:** Replacing SB with moderate or higher intensity PA may decrease AF risk. In REGARDS, these substitutions appeared to benefit older individuals and those reporting shorter sleep duration. These findings highlight the potential benefit of reducing sedentarism to lower AF burden, warranting validation in future studies.

## BACKGROUND

Atrial fibrillation (AF) is the most common sustained cardiac arrhythmia. In 2021, North America and Western Europe had the highest age-standardized prevalence of AF worldwide.^1^ Across community-based cohorts, lifetime AF risk has been estimated to be substantial in both men and women and across racial groups, although risk estimates and detection patterns may differ by population.^2–4^ People with AF are at increased risk of serious complications, including ischemic stroke, heart failure (HF), myocardial infarction (MI), cognitive impairment and dementia, and mortality.^2,3,5^ Despite this burden, effective detection and prevention strategies for AF are lacking, partially due to an incomplete comprehension of the etiopathogenesis and pathophysiology of AF. Moreover, although AF treatment has been identified as a major research priority, comparatively less attention has been paid towards developing effective, substantial approaches for prevention.^6^

Sedentary behavior is defined as any waking behavior characterized by an energy expenditure ≤1.5 metabolic equivalents (METs), while in a sitting, reclining, or lying posture.^7^ From 2002 to 2017, the prevalence of sedentary behavior, defined as ≥ 4.5 hours of sitting time per day, increased from 49.3% to 54.3% among European adults.^8,9^ In the United States, adults spend an average of 9.5 hours per day sedentary, with leisure activities accounting for nearly half of total sedentary time (47%).^10^ Evidence has linked sedentarism with an increased risk of CVDs and CVD mortality;^11–13^ and suggested that sedentary behavior is a risk factor for CVDs distinct from physical activity.^14–16^ However, “sedentary behavior” is still often used interchangeably with “physical inactivity,” terms that are conceptually distinct and should be applied consistently across studies. Sedentary behavior is also distinct from sleep, as it refers to specific low-energy-expenditure behaviors while awake. A 24-hour day comprises mutually exclusive periods of sedentary behavior, physical activity, and sleep; therefore, reducing sedentary behavior necessarily reallocates time to another behavior, and the health association may depend on the replacement behavior. Understanding whether AF risk differs when sedentary time is reallocated to physical activity or sleep duration can help inform public health messaging and clinical counseling. Examining this question across populations with different age structures, racial composition, baseline cardiometabolic risk, and movement behavior profiles may help assess the consistency and generalizability of findings. The UK Biobank (UKB) study and the REasons for Geographic And Racial Differences in Stroke study (REGARDS) provide complementary cohorts from the United Kingdom and the United States with objectively measured movement and non-movement behaviors, allowing the association between sedentary behavior reallocation and AF risk to be evaluated across two populations with high AF burden and different demographic and cardiometabolic characteristics.

## METHODS

### Study design and population

#### UK Biobank

UKB is a large, detailed prospective study of the middle- and old-age population in the United Kingdom. The primary purpose of the UKB is to investigate the genetic and nongenetic determinants of a wide range of diseases. Over 500,000 participants were recruited from 22 centers across the country at baseline from 2006 to 2010. Comprehensive data were collected through questionnaires, physical measures, imaging, sample assays, genome-wide genotyping at baseline. Longitudinal follow-up for health-related outcomes is ongoing. A detailed UKB cohort profile has been published previously.^17^ Participants who provided valid email addresses were invited to wear an accelerometer for a week between February 2013 and December 2015.^18^ Among those being invited, about half consented, and 45% sent back their devices.^18^ After excluding those without good wearing time (at least 72 hours of data and also having data in each one-hour period of the 24-hour cycle) and data quality calibration, and those who had AF before wearing the accelerometer, 86,096 provided valid data for the analysis. Figure 1a shows the flowchart of the study sample in this study. National electronic health-related data sets are linked to participants for health outcomes during follow-up.^17^ The National Health Service incorporated data from the national death registry, cancer registry, primary care record, and hospital inpatient records.^19^ This study is fully compliant with UK Biobank’s data access and ethical requirements. The research was conducted using data from the UK Biobank Resource under application number 34031.

**Figure 1.**
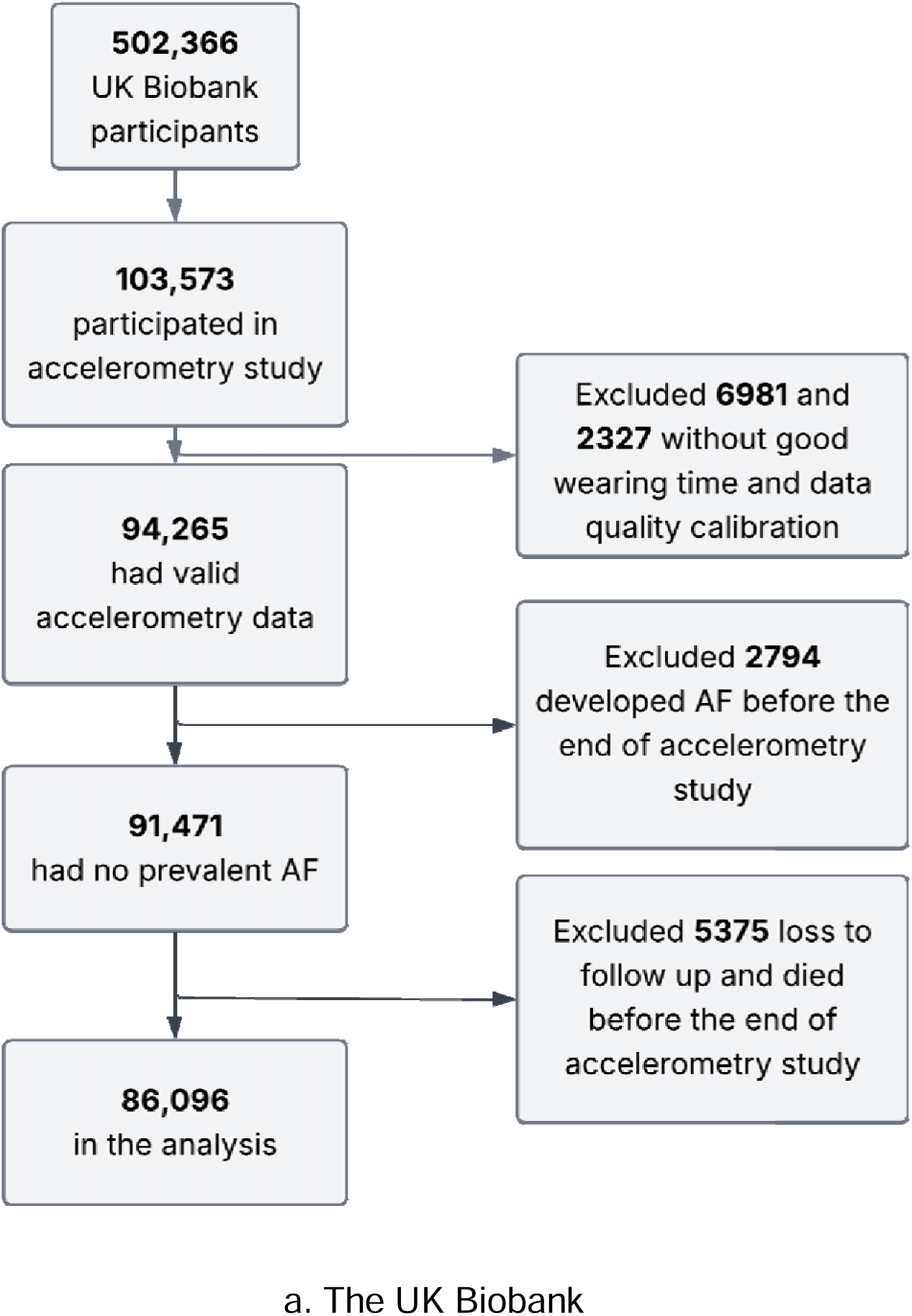
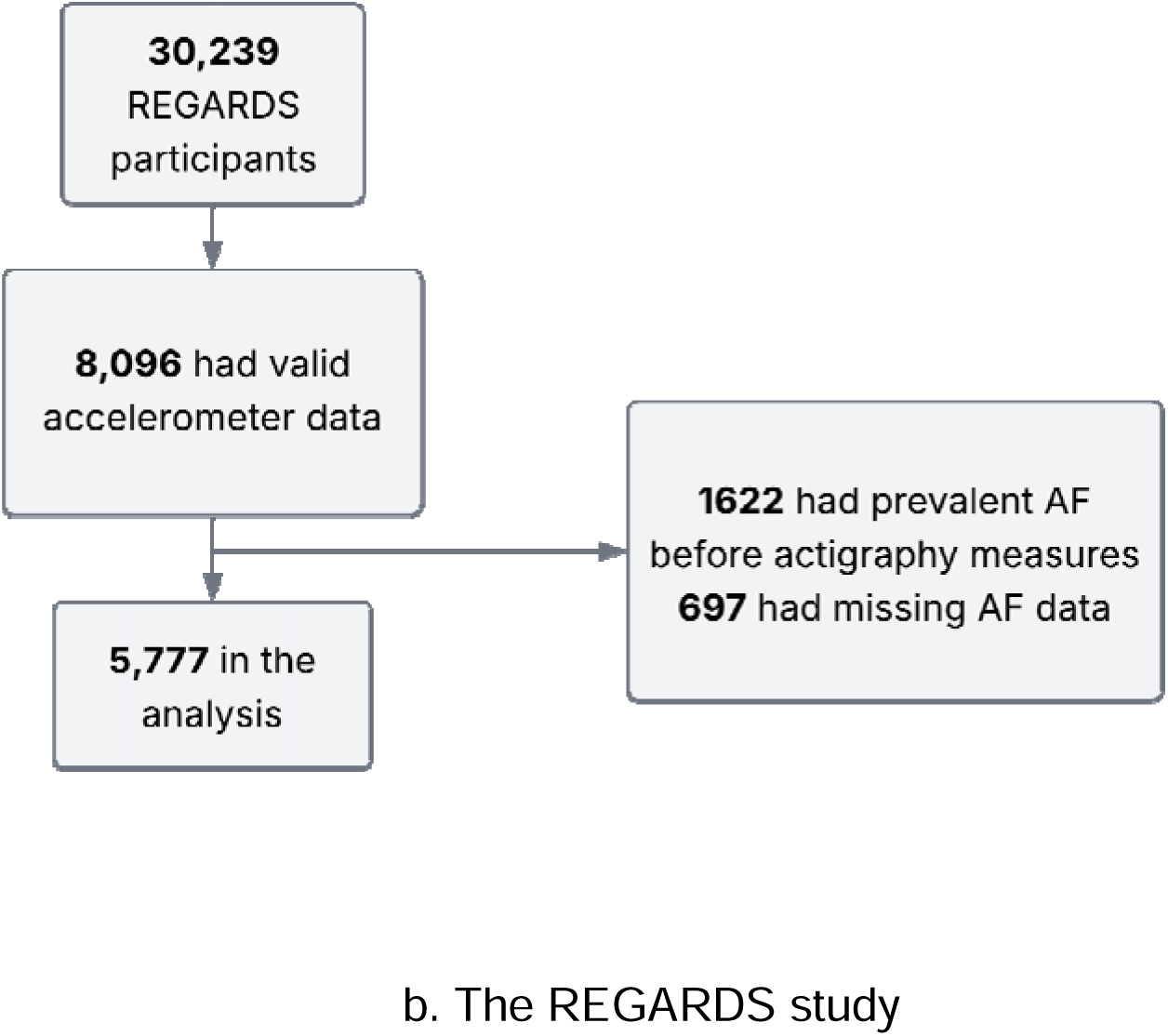
Flowchart of the study samples in UKB and REGARDS

#### REGARDS

The REGARDS study is a national longitudinal cohort study of non-Hispanic White and African American individuals in the US, ages 45 and older, with oversampling from the Stroke Belt in the Southeast US. The objective of the REGARDS study is to investigate the determinants of the excess stroke mortality in the Southeastern US and among African American individuals. During 2003-2007, selected from a commercially available nationwide list, participants were randomly recruited by mail and then verbal consent was obtained by computer-assisted telephone interview (CATI) during which data on sociodemographic, lifestyle, and psychosocial characteristics were collected. An in-home examination was carried out 3-4 weeks after the enrollment and written informed consent was obtained. Physical and physiological measures, a resting electrocardiogram (ECG), and fasting blood/urine samples were collected during the examination. The participants or their proxies were telephoned every 6 months, and potential stroke outcomes were adjudicated using medical records and/or death certificates. Detailed information on the REGARDS objectives and design was reported previously.^20^ A second CATI and in-home assessment similar to baseline was conducted approximately ten years later (2013-2016). Objective measures of sedentary behavior and physical activity by accelerometer were conducted between May 2009 - January 2013 in an ancillary study.^21^ Of the original cohort, 20,076 participants were invited to join the ancillary study of accelerometer and 12,146 (60.5%) consented and 9,973 (82.1%) wore and returned their devices. Participation rates were not significantly different by race-sex groups.^21^ Those who agreed to join had higher education levels and higher household income, and other differences were not clinically meaningful. Excluding those with device errors, missing or invalid data (wear time < 4 days with ≥ 10 hr/day, 8,096 had usable accelerometer data. Participants with AF before wearing the accelerometer or with missing AF data in the baseline assessment interview were also excluded. The study sample had a final size of 5,777 (Figure 1b). The REGARDS study was approved by the institutional review boards of participating institutions.

### Exposure measures

In the UKB, daily movement behaviors were measured using Axivity AX3, a wrist-worn triaxial accelerometer worn continuously on the dominant wrist for seven consecutive days. Accelerometer data were processed using a machine-learning model developed in the CAPTURE-24 study, which classified behaviors as sleep duration, sedentary behavior, light physical activity (LPA), and moderate-to-vigorous physical activity (MVPA) using accelerometer signals together with camera images and diary data.^22^ In REGARDS, an Actical accelerometer was used to collect data in 60-second epochs. Participants were instructed to wear the accelerometer snugly over the right hip for 7 days during waking hours and remove it at bedtime. Sedentary behavior, LPA, and MVPA were derived from accelerometry data, and the proportions of the time spent in sedentary behavior, LPA, and MVPA were calculated relative to total accelerometer wearing time.^23,24^ Sleep duration in REGARDS was self-reported in hours per day through CATI from 2008 to 2010 and was used for descriptive and stratified analyses rather than as an accelerometer-derived behavior component. In the present study, sleep duration was categorized as <7 versus ≥7 hours/day for stratified analyses based on the National Sleep Foundation sleep recommendation for adults.^25^

### Outcome measures

Incident AF is the primary outcome. In UKB, incident AF was classified by at least 1 ICD-10 code for the condition listed as either a primary diagnosis, secondary diagnosis, or cause of death through hospital and death registry data linkage. Incident AF included hospital diagnosed AF or death due to AF or flutter (I48.x).^26^ In REGARDS, incident AF was identified by the ECG during the second in-home assessment in 2013 to 2016, and by self-reported medical history during the CATI, based on an affirmative response to the question “Has a physician or a health professional ever told you that you had atrial fibrillation?”^27^

### Covariates

Covariates were selected a priori based on potential confounding and availability in each cohort and were assessed before the accelerometer assessment. In UKB, the minimally adjusted model included age, sex, ethnicity, assessment center, and accelerometer wear time. The fully adjusted model additionally included body mass index (BMI), systolic and diastolic blood pressure (SBP and DBP), waist circumference, total cholesterol, education, alcohol use, smoking, Townsend deprivation index, household income, medical history (heart attack, stroke, high blood pressure, deep vein thrombosis, pulmonary embolism, cancer, diabetes, bone fracture within the past 5 years, and other disability diagnoses), and use of cholesterol-lowering, blood pressure-lowering, and insulin medications. In REGARDS, the minimally adjusted model included age, sex, race, and accelerometer wear time; the fully adjusted model additionally included BMI, HDL and LDL cholesterol, systolic and diastolic blood pressure, education, alcohol use, smoking, marital/relationship status, and medical history of hypertension, diabetes, and MI. Because variable availability and coding differed between cohorts, cohort-specific adjustment sets were used rather than forcing identical covariates across studies. UKB covariates were obtained from questionnaires, physical measurements, and laboratory data collected at baseline.^28^ REGARDS covariates were obtained from the first CATI and in-home examination.^20^

### Statistical analysis

In both UKB and REGARDS, the study participants’ characteristics at baseline were described stratified by the tertiles of sedentary behavior. Mean (standard deviation [SD]) and frequency (percentage) were reported for continuous and categorical variables, respectively. Isotemporal substitution was used to examine the association between replacing 30 minutes of sedentary behavior with LPA, MVPA, and, in UKB, sleep duration at baseline and AF risk using Cox proportional hazards models in UKB and logistic regression in REGARDS. Model 1 is minimally adjusted and model 2 further adjusted for additional covariates. In UKB, the follow-up time was defined as time to incident AF, death, loss to follow-up, or censoring date (December 31, 2022 for England, August 31, 2022 for Scotland, and May 31, 2022 for Wales), whichever was the earliest. The proportional hazards assumption was tested. Additionally, the effect of sedentary behavior substitution was examined across baseline sedentary behavior tertiles and across categories defined by meeting the MVPA guideline of 150 minutes per week (yes vs. no). Stratified analyses across sex, age (<65 years vs. ≥65 years), and sleep hours (<7 hours vs. ≥7 hours) were performed and the interactions were tested. Hazard ratios (HRs) and their 95% CIs were reported. In REGARDS, replacing each 1% sedentary time of accelerometer wearing time with the same percentage of LPA and MVPA was also examined, in addition to 30-minute replacement. Odds ratios (ORs) and their 95% confidence intervals (CIs) were reported. In addition, stratified analyses were performed by sex, age (<65 years vs. ≥65 years), race (African American vs. White adults), and sleep duration (<7 hours vs. ≥7 hours), and interactions were tested using cross-product terms, with a two-sided p-value <0.05 considered statistically significant.

## RESULTS

### UKB

The study population’s mean age was 56.0 (SD 7.8) years, 57% were female, and 97.8% were White adults (n=86,096). Over a median follow-up time of 7.9 (interquartile range [IQR] 7.3-8.4) years, 4,035 incident AF cases (4.7%) were identified. The AF incident rate was 6.1 per 1,000 person-years among the full cohort and the rates increased across sedentary behavior tertiles, from 5.1 to 7.1 per 1,000 person-years. On average, participants spent 564 (SD 109) minutes (9.4 hours) per day being sedentary, 303 (SD 98) minutes (5.1 hours) in LPA, 42 (SD 35) minutes (0.7 hours) in MVPA, and 531 (SD 74) minutes (8.9 hours) sleeping (Table 1a). Across the sedentary behavior tertiles, participants tended to have higher BMI, blood pressure and education, lower overall health, and to be current smokers as the sedentary time increased. In the minimally adjusted model 1, substituting 30 minutes of sedentary behavior with LPA and MVPA was associated with a lower risk of AF (LPA: HR 0.98, 95% CI 0.97-0.99; MVPA HR 0.90, 95% CI 0.87-0.92) (Table 2). However, associations were attenuated and became non-significant after further adjusting for covariates in model 2 (LPA: HR 0.99, 95% CI 0.95-1.02; MVPA HR 1.00, 95% CI 0.99-1.02). In sex- and age-stratified analyses, substitution of sedentary behavior with MVPA appeared slightly protective in females and participants aged <65 years, but not in males or older adults (fully adjusted p-values for interaction 0.092 and 0.174). No effect was observed for substituting sedentary behavior with sleep in the full cohort, and across strata of sex or age, or across sleep-duration strata (Table 2). Table 3 shows the effect of sedentary behavior substitution across the baseline sedentary tertiles. In model 1, the HRs of MVPA substitution became gradually smaller with higher tertiles; In model 2, HRs in all the tertiles were close to null, and no monotonic pattern was present. Among those who did not meet the MVPA guideline, substituting sedentary behavior with MVPA tended to be associated with lower AF risk overall and across sedentary tertiles, although not statistically significant and the CIs were wide; among those meeting the MVPA guidelines, estimates were near null overall and in lower tertiles and shifted above null in the highest sedentary tertile without statistical significance (Figure 2).

**Figure 2.**
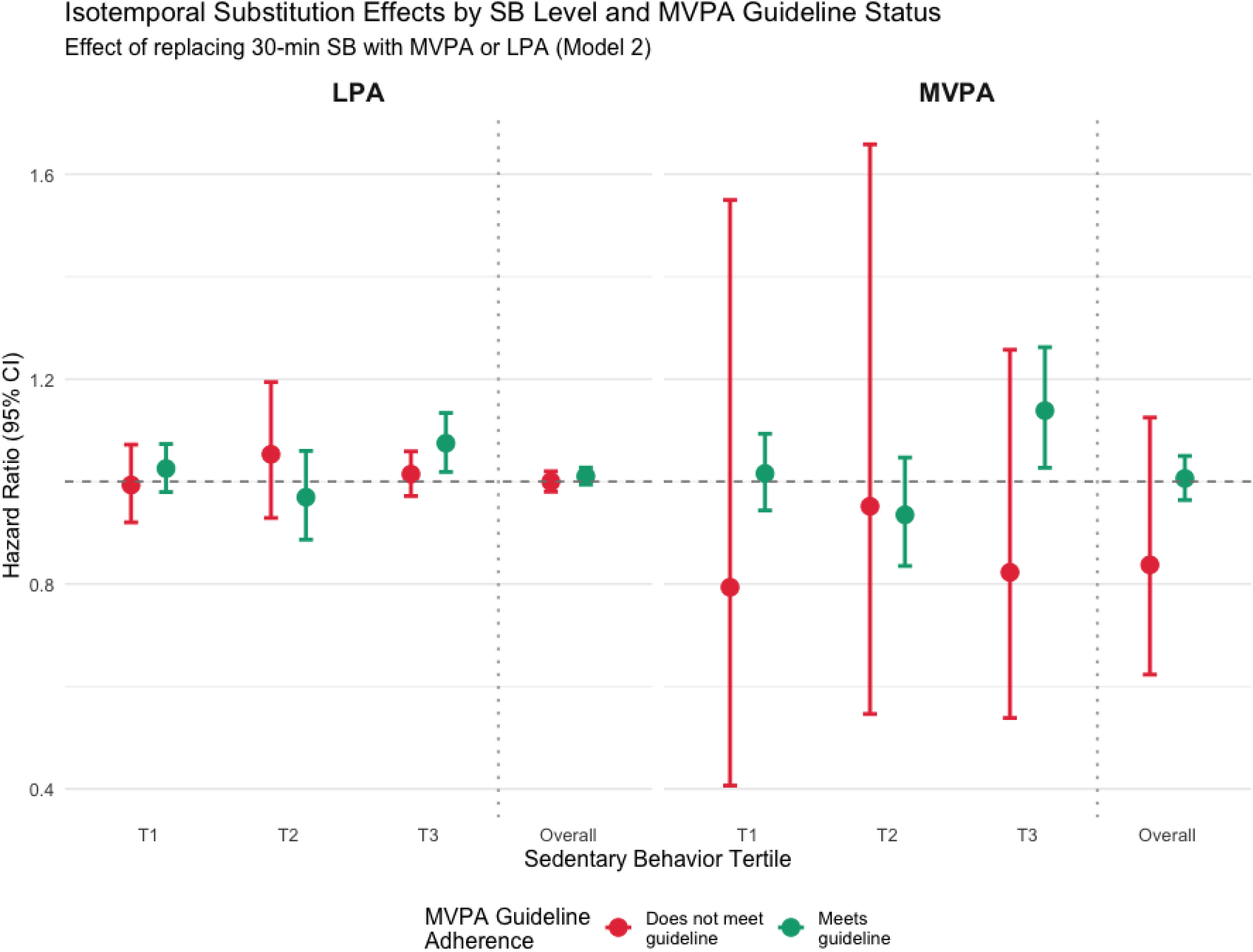
Isotemporal substitution of sedentary behavior with physical activities across sedentary behavior tertiles and the MVPA guideline, UK Biobank

**Table 1a.**
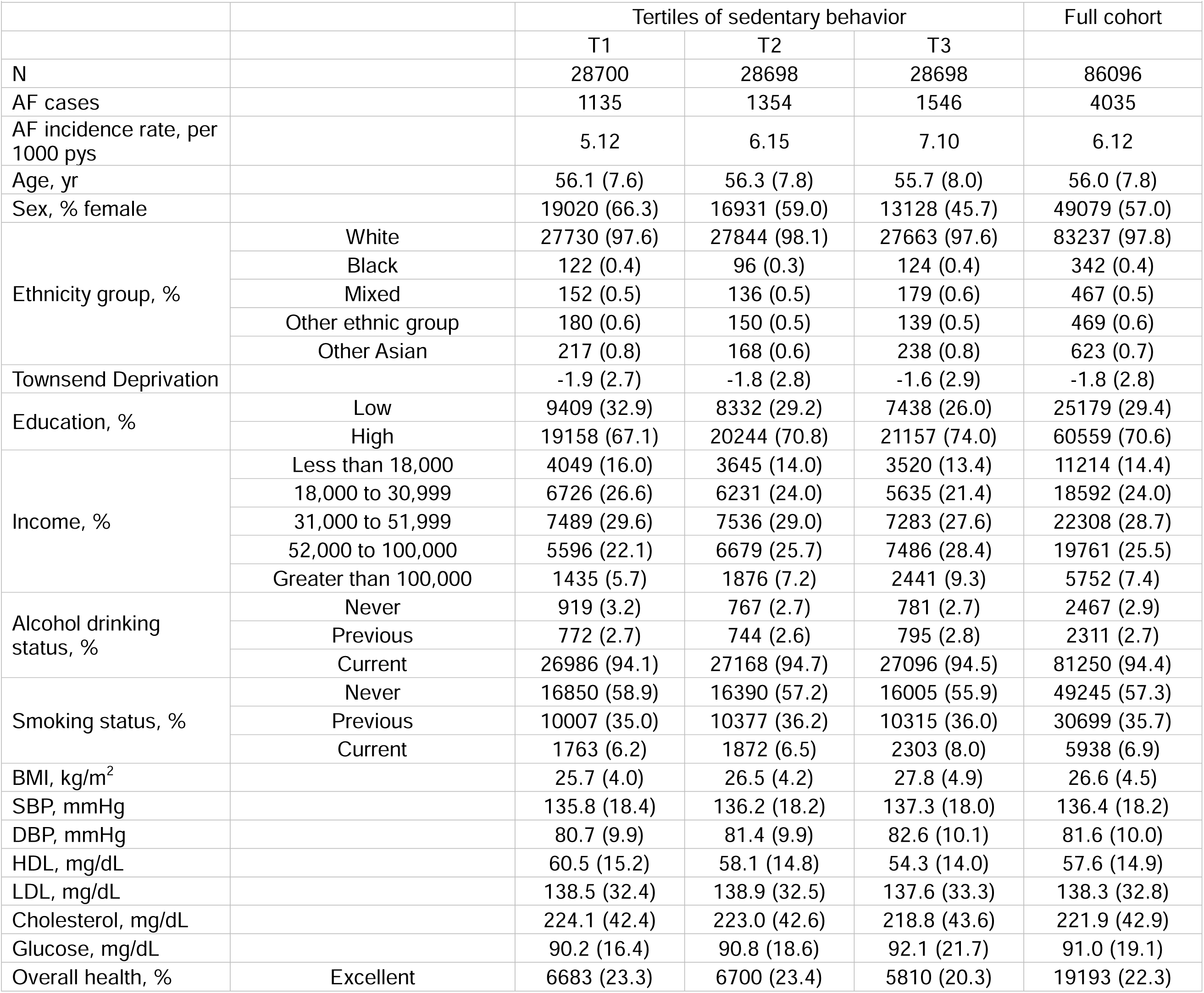
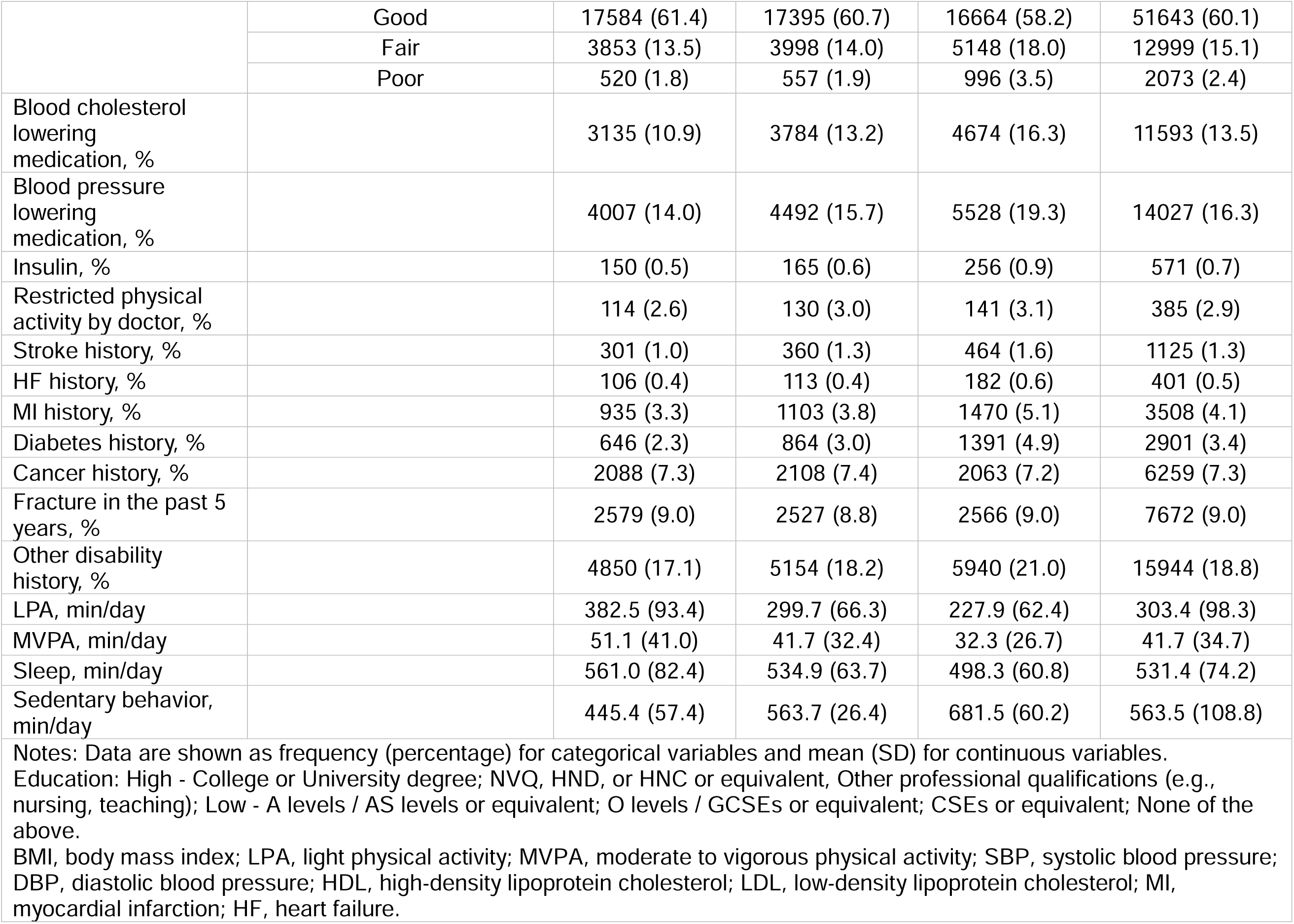
Baseline characteristics of participants in the UK Biobank.

| Table 1a. Baseline characteristics of participants in the UK Biobank |  |  |  |  |  |
| --- | --- | --- | --- | --- | --- |
|  |  | Tertiles of sedentary behavior |  |  | Full cohort |
|  |  | T1 | T2 | T3 |  |
| N |  | 28700 | 28698 | 28698 | 86096 |
| AF cases |  | 1135 | 1354 | 1546 | 4035 |
| AF incidence rate, per 1000 pys |  | 5.12 | 6.15 | 7.10 | 6.12 |
| Age, yr |  | 56.1 (7.6) | 56.3 (7.8) | 55.7 (8.0) | 56.0 (7.8) |
| Sex, % female |  | 19020 (66.3) | 16931 (59.0) | 13128 (45.7) | 49079 (57.0) |
| Ethnicity group, % | White | 27730 (97.6) | 27844 (98.1) | 27663 (97.6) | 83237 (97.8) |
|  | Black | 122 (0.4) | 96 (0.3) | 124 (0.4) | 342 (0.4) |
|  | Mixed | 152 (0.5) | 136 (0.5) | 179 (0.6) | 467 (0.5) |
|  | Other ethnic group | 180 (0.6) | 150 (0.5) | 139 (0.5) | 469 (0.6) |
|  | Other Asian | 217 (0.8) | 168 (0.6) | 238 (0.8) | 623 (0.7) |
| Townsend Deprivation |  | -1.9 (2.7) | -1.8 (2.8) | -1.6 (2.9) | -1.8 (2.8) |
| Education, % | Low | 9409 (32.9) | 8332 (29.2) | 7438 (26.0) | 25179 (29.4) |
|  | High | 19158 (67.1) | 20244 (70.8) | 21157 (74.0) | 60559 (70.6) |
| Income, % | Less than 18,000 | 4049 (16.0) | 3645 (14.0) | 3520 (13.4) | 11214 (14.4) |
|  | 18,000 to 30,999 | 6726 (26.6) | 6231 (24.0) | 5635 (21.4) | 18592 (24.0) |
|  | 31,000 to 51,999 | 7489 (29.6) | 7536 (29.0) | 7283 (27.6) | 22308 (28.7) |
|  | 52,000 to 100,000 | 5596 (22.1) | 6679 (25.7) | 7486 (28.4) | 19761 (25.5) |
|  | Greater than 100,000 | 1435 (5.7) | 1876 (7.2) | 2441 (9.3) | 5752 (7.4) |
| Alcohol drinking status, % | Never | 919 (3.2) | 767 (2.7) | 781 (2.7) | 2467 (2.9) |
|  | Previous | 772 (2.7) | 744 (2.6) | 795 (2.8) | 2311 (2.7) |
|  | Current | 26986 (94.1) | 27168 (94.7) | 27096 (94.5) | 81250 (94.4) |
| Smoking status, % | Never | 16850 (58.9) | 16390 (57.2) | 16005 (55.9) | 49245 (57.3) |
|  | Previous | 10007 (35.0) | 10377 (36.2) | 10315 (36.0) | 30699 (35.7) |
|  | Current | 1763 (6.2) | 1872 (6.5) | 2303 (8.0) | 5938 (6.9) |
| BMI, kg/m<sup>2</sup> |  | 25.7 (4.0) | 26.5 (4.2) | 27.8 (4.9) | 26.6 (4.5) |
| SBP, mmHg |  | 135.8 (18.4) | 136.2 (18.2) | 137.3 (18.0) | 136.4 (18.2) |
| DBP, mmHg |  | 80.7 (9.9) | 81.4 (9.9) | 82.6 (10.1) | 81.6 (10.0) |
| HDL, mg/dL |  | 60.5 (15.2) | 58.1 (14.8) | 54.3 (14.0) | 57.6 (14.9) |
| LDL, mg/dL |  | 138.5 (32.4) | 138.9 (32.5) | 137.6 (33.3) | 138.3 (32.8) |
| Cholesterol, mg/dL |  | 224.1 (42.4) | 223.0 (42.6) | 218.8 (43.6) | 221.9 (42.9) |
| Glucose, mg/dL |  | 90.2 (16.4) | 90.8 (18.6) | 92.1 (21.7) | 91.0 (19.1) |
| Overall health, % | Excellent | 6683 (23.3) | 6700 (23.4) | 5810 (20.3) | 19193 (22.3) |
|  | Good | 17584 (61.4) | 17395 (60.7) | 16664 (58.2) | 51643 (60.1) |
|  | Fair | 3853 (13.5) | 3998 (14.0) | 5148 (18.0) | 12999 (15.1) |
|  | Poor | 520 (1.8) | 557 (1.9) | 996 (3.5) | 2073 (2.4) |
| Blood cholesterol lowering medication, % |  | 3135 (10.9) | 3784 (13.2) | 4674 (16.3) | 11593 (13.5) |
| Blood pressure lowering medication, % |  | 4007 (14.0) | 4492 (15.7) | 5528 (19.3) | 14027 (16.3) |
| Insulin, % |  | 150 (0.5) | 165 (0.6) | 256 (0.9) | 571 (0.7) |
| Restricted physical activity by doctor, % |  | 114 (2.6) | 130 (3.0) | 141 (3.1) | 385 (2.9) |
| Stroke history, % |  | 301 (1.0) | 360 (1.3) | 464 (1.6) | 1125 (1.3) |
| HF history, % |  | 106 (0.4) | 113 (0.4) | 182 (0.6) | 401 (0.5) |
| MI history, % |  | 935 (3.3) | 1103 (3.8) | 1470 (5.1) | 3508 (4.1) |
| Diabetes history, % |  | 646 (2.3) | 864 (3.0) | 1391 (4.9) | 2901 (3.4) |
| Cancer history, % |  | 2088 (7.3) | 2108 (7.4) | 2063 (7.2) | 6259 (7.3) |
| Fracture in the past 5 years, % |  | 2579 (9.0) | 2527 (8.8) | 2566 (9.0) | 7672 (9.0) |
| Other disability history, % |  | 4850 (17.1) | 5154 (18.2) | 5940 (21.0) | 15944 (18.8) |
| LPA, min/day |  | 382.5 (93.4) | 299.7 (66.3) | 227.9 (62.4) | 303.4 (98.3) |
| MVPA, min/day |  | 51.1 (41.0) | 41.7 (32.4) | 32.3 (26.7) | 41.7 (34.7) |
| Sleep, min/day |  | 561.0 (82.4) | 534.9 (63.7) | 498.3 (60.8) | 531.4 (74.2) |
| Sedentary behavior, min/day |  | 445.4 (57.4) | 563.7 (26.4) | 681.5 (60.2) | 563.5 (108.8) |
| Notes: Data are shown as frequency (percentage) for categorical variables and mean (SD) for continuous variables.<br>Education: High - College or University degree; NVQ, HND, or HNC or equivalent, Other professional qualifications (e.g., nursing, teaching); Low - A levels / AS levels or equivalent; O levels / GCSEs or equivalent; CSEs or equivalent; None of the above.<br>BMI, body mass index; LPA, light physical activity; MVPA, moderate to vigorous physical activity; SBP, systolic blood pressure; DBP, diastolic blood pressure; HDL, high-density lipoprotein cholesterol; LDL, low-density lipoprotein cholesterol; MI, myocardial infarction; HF, heart failure. |  |  |  |  |  |

**Table 1b.** Baseline characteristics of participants in the REGARDS study.

|  |  |  |  |  |  |
| --- | --- | --- | --- | --- | --- |
|  |  | Tertiles of sedentary behavior |  |  | Full cohort |
|  |  | T1 | T2 | T3 |  |
| N |  | 1925 | 1927 | 1925 | 5777 |
| AF cases |  | 129 | 160 | 197 | 486 |
| Age, yr |  | 60.6 (7.5) | 63.1 (8.0) | 65.2 (8.4) | 63.0 (8.2) |
| Sex, female, % |  | 1055 (54.8) | 1085 (56.3) | 990 (51.4) | 3130 (54.2) |
| Race, black, % |  | 573 (29.8) | 518 (26.9) | 671 (34.9) | 1762 (30.5) |
| BMI, kg/m<sup>2</sup> |  | 28.1 (5.1) | 28.6 (5.7) | 29.2 (5.9) | 28.7 (5.6) |
| Education, % | Less than high school | 112 (5.8) | 84 (4.4) | 110 (5.7) | 306 (5.3) |
|  | High school graduate | 420 (21.8) | 405 (21) | 398 (20.7) | 1223 (21.2) |
|  | Some college | 539 (28) | 498 (25.8) | 492 (25.6) | 1529 (26.5) |
|  | College graduate and above | 854 (44.4) | 940 (48.8) | 924 (48) | 2718 (47.1) |
| Alcohol use, % | Current | 1209 (62.8) | 1186 (61.6) | 1110 (57.7) | 3505 (60.7) |
|  | Past | 249 (12.9) | 240 (12.5) | 289 (15) | 778 (13.5) |
|  | Never | 467 (24.3) | 501 (26) | 526 (27.3) | 1494 (25.9) |
| Smoking, % | Current | 219 (11.4) | 171 (8.9) | 169 (8.8) | 559 (9.7) |
|  | Past | 772 (40.2) | 734 (38.3) | 804 (41.9) | 2310 (40.1) |
|  | Never | 467 (24.3) | 501 (26) | 526 (27.3) | 1494 (25.9) |
| Relationship status, % | Divorced | 227 (11.8) | 226 (11.7) | 270 (14) | 723 (12.5) |
|  | Married | 1382 (71.8) | 1324 (68.7) | 1233 (64.1) | 3939 (68.2) |
|  | Other | 34 (1.8) | 28 (1.5) | 30 (1.6) | 92 (1.6) |
|  | Single | 99 (5.1) | 93 (4.8) | 72 (3.7) | 264 (4.6) |
|  | Widowed | 183 (9.5) | 256 (13.3) | 320 (16.6) | 759 (13.1) |
| SBP, mmHg |  | 123.6 (15.3) | 124.3 (15.0) | 126.3 (15.3) | 124.7 (15.2) |
| DBP, mmHg |  | 76.3 (9.0) | 75.6 (9.0) | 76.3 (9.4) | 76.1 (9.1) |
| HDL, mg/dL |  | 54.1 (16.8) | 53.1 (16.0) | 51.7 (15.9) | 52.9 (16.3) |
| LDL, mg/dL |  | 117.0 (34.4) | 115.0 (32.6) | 113.3 (33.0) | 115.1 (33.4) |
| Hypertension, % |  | 1209 (62.8) | 1268 (66.0) | 1394 (72.6) | 3871 (67.2) |
| Diabetes, % |  | 185 (9.8) | 256 (13.8) | 316 (16.9) | 757 (13.5) |
| MI, % |  | 109 (5.7) | 142 (7.5) | 156 (8.2) | 407 (7.2) |
| LPA, minutes/day |  | 245.1 (73.1) | 189.6 (60.7) | 147.9 (61.4) | 194.2 (76.5) |
| MPA, minuets/day |  | 20.5 (21.7) | 13.3 (16.0) | 8.9 (12.5) | 14.2 (17.8) |
| VPA, minutes/day |  | 0.4 (2.5) | 0.2 (1.6) | 0.1 (1.3) | 0.2 (1.9) |
| MVPA,<br>minutes/day |  | 20.8 (22.3) | 13.5 (16.3) | 9.0 (12.8) | 14.5 (18.2) |
| Sedentary<br>behavior,<br>minutes/day |  | 570.9 (55.3) | 681.1 (24.5) | 806.2 (90.3) | 686.1 (114.8) |
| Sleep, hours/day |  | 8.1 (2.6) | 8.1 (2.8) | 8.0 (3.0) | 8.1 (2.8) |
| Notes: Data are shown as frequency (percentage) for categorical variables and mean (SD) for continuous variables.<br>BMI, body mass index; LPA, light physical activity; MPA, moderate physical activity; VPA, vigorous physical activity; MVPA, moderate to vigorous physical activity; SBP, systolic blood pressure; DBP, diastolic blood pressure; HDL, high-density lipoprotein cholesterol; LDL, low-density lipoprotein cholesterol; eGFR, estimated glomerular filtration rate; CRP, c-reactive protein; NT-proBNP, N-terminal pro-B-type natriuretic peptide; MI, myocardial infarction. |  |  |  |  |  |

**Table 2.** Isotemporal substitution of sedentary time (30min/day) and sedentary percentage with physical activities on AF risk (hazard ratios), UK Biobank.

|  | LPA |  | MVPA |  | Sleep |  |
| --- | --- | --- | --- | --- | --- | --- |
|  | Estimate (95% CI) | p value for interaction | Estimate (95% CI) | p value for interaction | Estimate (95% CI) | p value for interaction |
| Model 1 | 0.98 (0.97, 0.99) |  | 0.90 (0.87, 0.92) |  | 0.99 (0.97, 1.00) |  |
| Model 2 | 0.99 (0.95, 1.02) |  | 1.00 (0.99, 1.02) |  | 0.99 (0.98, 1.01) |  |
| **Male** |  |  |  |  |  |  |
| Model 1 | 1.00 (0.98, 1.01) | 0.003 | 0.91 (0.88, 0.94) | 0.029 | 0.99 (0.97, 1.01) | 0.642 |
| Model 2 | 1.02 (1.00, 1.03) | 0.010 | 0.99 (0.96, 1.03) | 0.092 | 1.00 (0.98, 1.02) | 0.469 |
| **Female** |  |  |  |  |  |  |
| Model 1 | 0.96 (0.94, 0.98) |  | 0.84 (0.80, 0.90) |  | 0.98 (0.96, 1.00) |  |
| Model 2 | 0.98 (0.96, 1.01) |  | 0.94 (0.88, 1.01) |  | 0.99 (0.96, 1.01) |  |
| **Age < 65** |  |  |  |  |  |  |
| Model 1 | 0.97 (0.96, 0.98) | 0.001 | 0.88 (0.84, 0.91) | 0.193 | 0.98 (0.96, 1.00) | 0.101 |
| Model 2 | 0.99 (0.98, 1.01) | 0.002 | 0.97 (0.93, 1.01) | 0.174 | 0.99 (0.97, 1.01) | 0.425 |
| **Age ≥ 65** |  |  |  |  |  |  |
| Model 1 | 1.00 (0.98, 1.02) |  | 0.93 (0.88, 0.98) |  | 1.00 (0.98, 1.02) |  |
| Model 2 | 1.03 (1.01, 1.05) |  | 1.02 (0.96, 1.08) |  | 1.00 (0.97, 1.02) |  |
| **Sleep ≥ 7 hours** |  |  |  |  |  |  |
| Model 1 | 0.98 (0.97, 0.99) | 0.352 | 0.90 (0.87, 0.93) | 0.530 | 0.99 (0.98, 1.01) | 0.254 |
| Model 2 | 1.00 (0.98, 1.01) | 0.190 | 1.01 (0.97, 1.04) | 0.523 | 1.00 (0.98, 1.01) | 0.269 |
| **Sleep < 7 hours** |  |  |  |  |  |  |
| Model 1 | 1.00 (0.97, 1.04) |  | 0.90 (0.80, 1.03) |  | 0.95 (0.86, 1.05) |  |
| Model 2 | 1.02 (0.98, 1.07) |  | 1.06 (0.94, 1.19) |  | 0.94 (0.85, 1.05) |  |

Notes: Cox proportional hazards models were used.
Model 1: adjusted for age, sex, ethnicity, assessment center
Model 2: model 1 + BMI, SBP, DBP, total cholesterol, waist, education, alcohol use, smoking, Townsend deprivation index, income, medical history (heart attack, stroke, high blood pressure, blood clot in the leg, blood clot in the lung, cancer, diabetes, bone fracture within 5 years, other disability diagnosis), medications (cholesterol, blood pressure, insulin).

**Table 3.** Isotemporal substitution of sedentary time (30min/day) with physical activities and sleep across tertiles of sedentary behavior on AF risk (hazard ratios), UK Biobank.

|  | SB<br>tertiles | LPA | MVPA | Sleep |
| --- | --- | --- | --- | --- |
| Model 1 | T1 | 1.00 (0.96, 1.03) | 0.93 (0.88, 0.98) | 1.02 (0.98, 1.06) |
|  | T2 | 0.98 (0.92, 1.05) | 0.88 (0.82, 0.96) | 0.98 (0.92, 1.05) |
|  | T3 | 0.99 (0.96, 1.02) | 0.88 (0.82, 0.94) | 0.97 (0.94, 1.00) |
| Model 2 | T1 | 1.01 (0.98, 1.06) | 0.99 (0.93, 1.05) | 1.02 (0.98, 1.06) |
|  | T2 | 1.00 (0.93, 1.07) | 0.96 (0.88, 1.05) | 0.99 (0.92, 1.06) |
|  | T3 | 1.03 (1.00, 1.06) | 1.02 (0.95, 1.10) | 0.98 (0.95, 1.02) |

Notes: Cox proportional hazards models were used.
Model 1: adjusted for age, sex, ethnicity, assessment center
Model 2: model 1 + BMI, SBP, DBP, total cholesterol, waist, education, alcohol use, smoking, Townsend deprivation index, income, medical history (heart attack, stroke, high blood pressure, blood clot in the leg, blood clot in the lung, cancer, diabetes, bone fracture within 5 years, other disability diagnosis), medications (cholesterol, blood pressure, insulin).

### REGARDS

The study participants (n=5,777) had a mean age of 63.0 (SD 8.3) years, 54% were female, and 31% were African American. The mean daily times spent on LPA, MVPA, and sedentary behavior were 194 (SD 76.5), 14.5 (SD 18.2), and 686.1 (SD 114.8) minutes, corresponding to 3.2, 0.2, and 11.4 hours; mean self-reported sleep duration was 8.1 (SD 2.8) hours/day. Participants were more likely to have higher BMI, hypertension, diabetes and MI history as sedentary behavior increased. During the follow-up exam after collecting the actigraphy data, 486 (8.4%) AF cases were recorded (Table 1b). Substituting 30 minutes of sedentary behavior per day with MVPA was associated with decreased odds of AF (OR 0.68, 95% CI 0.54, 0.86) after minimal adjustment, and the association was attenuated after further adjusting for covariates (OR 0.80, 95% CI 0.63-1.01), whereas LPA showed no association (Table 4). Similarly, replacing 1% of sedentary time with MVPA was linked to decreased odds of AF in both models (model 1: OR 0.89, 95% CI 0.83-0.96; model 2: OR 0.94, 95% CI 0.87-1.00), but no association was observed for LPA. The association between MVPA substitution and AF was more pronounced among females than males, although the interaction did not suggest significant difference. Among those aged ≥65 years, replacing sedentary behavior with MVPA was associated with lower AF odds in both 30 minutes (model 2: OR 0.64, 95% CI 0.44-0.93) and 1% reallocation analyses (model 2: OR 0.87, 95% CI 0.78-0.98). In the <65 years group, the corresponding MVPA estimates were in the same direction but attenuated after adjustment and were not statistically significant. In race-stratified analyses, point estimates for 30-minute MVPA substitution were nearly identical among White and Black participants, but the estimate among Black participants was substantially less precise (White: OR 0.81, 95% CI 0.63-1.04; Black: OR 0.81, 95% CI 0.40-1.62), with no evidence of interaction. By sleep duration, the association between MVPA substitution and AF was stronger among participants reporting <7 hours/day of sleep (30-minute OR 0.57, 95% CI 0.32-1.00; 1% OR 0.84, 95% CI 0.70-1.00) than among those reporting ≥7 hours/day (30-minute OR 0.87, 95% CI 0.68-1.12; 1% OR 0.96, 95% CI 0.89-1.04), with evidence of interaction for MVPA substitution (p-value 0.050 for 30-minute reallocation and 0.038 for 1% reallocation; Table 4).

**Table 4.** Isotemporal substitution of sedentary time (30min/day) and sedentary percentage with physical activities on AF risk (odds ratios), REGARDS study.

|  | Sedentary time (30 min) |  |  |  | Sedentary percentage (each 1%) |  |  |  |
| --- | --- | --- | --- | --- | --- | --- | --- | --- |
|  | LPA | p value for interaction | MVPA | p value for interaction | LPA | p value for interaction | MVPA | p value for interaction |
| Model 1 | 0.97 (0.92, 1.01) |  | 0.68 (0.54, 0.86) |  | 0.99 (0.98, 1.00) |  | 0.89 (0.83, 0.96) |  |
| Model 2 | 1.00 (0.95, 1.05) |  | 0.80 (0.63, 1.01) |  | 1.00 (0.99, 1.02) |  | 0.94 (0.87, 1.00) |  |
| **Male** |  |  |  |  |  |  |  |  |
| Model 1 | 0.98 (0.92, 1.04) | 0.285 | 0.73 (0.56, 0.95) | 0.104 | 0.99 (0.98, 1.01) | 0.343 | 0.91 (0.85, 0.99) | 0.072 |
| Model 2 | 1.02 (0.95, 1.08) | 0.382 | 0.87 (0.66, 1.13) | 0.183 | 1.01 (0.99, 1.03) | 0.407 | 0.96 (0.89, 1.04) | 0.146 |
| **Female** |  |  |  |  |  |  |  |  |
| Model 1 | 0.95 (0.89, 1.03) |  | 0.50 (0.30, 0.81) |  | 0.99 (0.97, 1.01) |  | 0.80 (0.69, 0.93) |  |
| Model 2 | 0.98 (0.90, 1.06) |  | 0.60 (0.36, 1.01) |  | 0.99 (0.97, 1.02) |  | 0.85 (0.73, 1.00) |  |
| **Age < 65** |  |  |  |  |  |  |  |  |
| Model 1 | 0.98 (0.91, 1.05) | 0.257 | 0.72 (0.54, 0.97) | 0.226 | 1.00 (0.98, 1.02) | 0.366 | 0.93 (0.85, 1.01) | 0.144 |
| Model 2 | 0.99 (0.92, 1.07) | 0.449 | 0.86 (0.63, 1.17) | 0.268 | 1.00 (0.98, 1.02) | 0.375 | 0.96 (0.87, 1.05) | 0.157 |
| **Age ≥ 65** |  |  |  |  |  |  |  |  |
| Model 1 | 0.92 (0.86, 0.98) |  | 0.55 (0.38, 0.80) |  | 0.99 (0.97, 1.00) |  | 0.84 (0.76, 0.94) |  |
| Model 2 | 0.96 (0.90, 1.02) |  | 0.64 (0.44, 0.93) |  | 0.99 (0.97, 1.01) |  | 0.87 (0.78, 0.98) |  |
| **White individuals** |  |  |  |  |  |  |  |  |
| Model 1 | 0.98 (0.93, 1.04) | 0.124 | 0.65 (0.51, 0.84) | 0.625 | 1.00 (0.98, 1.01) | 0.252 | 0.88 (0.82, 0.95) | 0.704 |
| Model 2 | 1.01 (0.96, 1.07) | 0.203 | 0.81 (0.63, 1.04) | 0.608 | 1.00 (0.99, 1.02) | 0.343 | 0.94 (0.87, 1.02) | 0.709 |
| **African Americans** |  |  |  |  |  |  |  |  |
| Model 1 | 0.90 (0.81, 1.01) |  | 0.81 (0.43, 1.54) |  | 0.97 (0.94, 1.00) |  | 0.94 (0.77, 1.14) |  |
| Model 2 | 0.94 (0.84, 1.06) |  | 0.81 (0.40, 1.62) |  | 0.99 (0.95, 1.02) |  | 0.94 (0.76, 1.15) |  |
| **Sleep ≥ 7 hours** |  |  |  |  |  |  |  |  |
| Model 1 | 0.97 (0.92, 1.03) | 0.572 | 0.77 (0.60, 0.99) | 0.036 | 0.99 (0.98, 1.01) | 0.828 | 0.92 (0.86, 1.00) | 0.022 |
| Model 2 | 1.01 (0.95, 1.07) | 0.425 | 0.87 (0.68, 1.12) | 0.050 | 1.00 (0.99, 1.02) | 0.653 | 0.96 (0.89, 1.04) | 0.038 |
| **Sleep < 7 hours** |  |  |  |  |  |  |  |  |
| Model 1 | 0.95 (0.86, 1.04) |  | 0.44 (0.26, 0.76) |  | 0.98 (0.96, 1.01) |  | 0.78 (0.66, 0.92 ) |  |
| Model 2 | 0.99 (0.90, 1.10) |  | 0.57 (0.32, 1.00) |  | 1.00 (0.97, 1.03) |  | 0.84 (0.70, 1.00) |  |

Notes: Logistic regression was used.
Model 1: adjusted for age, sex, race, total wearing time
Model 2: model 1 + BMI, education, alcohol use, smoking, relationship, hypertension, diabetes, MI, HDL, LDL, SBP, DBP
Sedentary, LPA and MVPA (or MPA and VPA) percentages were calculated as time/accelerometer wearing time. Participants were asked to take the accelerometer off during sleep.

## DISCUSSION

In this analysis of two large prospective cohorts with accelerometer-assessed movement behavior in the UK and the US, replacing sedentary behavior with MVPA was associated with lower AF risk in minimally adjusted models. After additional covariate adjustment, the estimates were attenuated, with MVPA substitution becoming statistically non-significant in UKB; in REGARDS, the association remained in the direction of lower AF odds, but its 95% CI included the null. LPA substitution showed little evidence of an independent effect on AF risk in either cohort. In UKB, no association was observed for substituting sedentary behavior with sleep, and sleep duration did not materially modify the associations of LPA or MVPA substitution with AF risk. In REGARDS, the inverse association between MVPA substitution and AF risk appeared stronger among participants aged ≥65 years and among those reporting <7 hours/day of sleep, while race-stratified estimates were imprecise and did not provide clear evidence of heterogeneity by African Americans versus White race.

Our findings align with and expand earlier work showing that greater sedentary time is associated with higher CVD risk, particularly incident AF. ^12,13,16,27,29–31^ A recent study utilizing accelerometer data in the UKB reported that higher sedentary time was associated with a greater risk of incident AF (HR 1.11, 95% CI 1.01-1.21) with a dose response, and reallocating sedentary time to MVPA substantially reduced the CV risk granted by sedentarism.^29^ Among older women in the US, prolonged sedentary time and patterns of sedentary bouts were both associated with a higher risk of incident AF, with estimates being attenuated after serial adjustment.^31^ In REGARDS, it was reported that objectively measured MVPA was beneficial for reducing the incident AF risk.^27^ These studies evaluated behavior-specific associations. In contrast, isotemporal substitution studies have examined time reallocation for broader cardiovascular outcomes. Walmsley et al. reported lower incident CVD risk when sedentary time was reallocated to MVPA in UKB;^22^ while Cao et al. used self-reported movement data in UKB to evaluate replacement of sedentary time across multiple adverse health outcomes.^30^ However, whether the association between replacing sedentary behavior with physical activity of different intensities and AF risk is consistent across populations with different sociodemographic characteristics and baseline AF risk remains unclear. Furthermore, self-reported activity data, a small sample size, and highly correlated movement behavior may limit the validity of these studies.

The protective effect observed in this study for MVPA substitution of sedentary behavior after multivariable adjustment in REGARDS is biologically plausible. Biological responses to sedentary behavior that may underline cardiovascular disease pathophysiology are likely multifactorial, involving peripheral organs crucial for metabolism, such as adipose tissue, the heart, and skeletal muscle. Mechanisms hypothesized to link sedentary behavior to cardiovascular health include various cardiometabolic risk factors, such as vascular structure and function, endothelial function, inflammation, insulin resistance, and blood lipids.^15,32^ Previous work found that prolonged, uninterrupted sitting was linked to increased hydrostatic pressure on the lower limbs, decreased shear rate in the popliteal artery, and impaired lower limb endothelial function, subsequently leading to vascular dysfunction.^33,34^ Longer sitting time was associated with elevated levels of adipokines, CRP, triglycerides, low-grade inflammation, and low density lipoprotein cholesterol.^35–37^ Replacing sedentary behavior with MVPA may reduce AF risk through improvements in cardiorespiratory fitness, cardiometabolic risk factors, autonomic balance, and weight regulation. It has been found that LPA and MVPA substitutions were beneficial for an improvement in cardiometabolic biomarkers, including CRP, glucose, and HDL-C.^38^ Although mechanisms linking sedentary behavior specifically to AF remain incompletely understood, our findings suggest that the intensity of the activity replacing sedentary time may be important.

The inclusion of both UKB and REGARDS strengthens this report by allowing the same substitution question to be evaluated in cohorts with different geographic settings, racial composition, age distribution, baseline cardiometabolic risk, accelerometer protocols, and AF ascertainment methods. These differences are also important for interpreting heterogeneity in the estimates. REGARDS participants were older, had a higher BMI on average, included a substantially larger proportion of African Americans, and accumulated less MVPA than UKB participants, such that replacing 30 minutes per day or 1% (approximately equivalent to 9 minutes) of accelerometer wearing time represented a larger relative movement change and possibly a greater physiological stimulus. In the subgroup of those who did not meet the MVPA guideline in UKB, the same direction of the association was observed as in REGARDS, although without statistical significance. In addition, the race and ethnicity distribution and baseline CVD risk may result in different estimates. A prior study suggested that “healthy volunteer” selection bias may exist in UKB.^39^ Moreover, the difference in AF ascertainment methods and follow-up structure could contribute to differences in the estimates. REGARDS identified incident AF cases at a follow-up exam after actigraphy measurement, and the incident date was not collected due to the design; UKB captured incident AF over a longer period of follow-up using linked medical records. Despite these differences, the consistent protective effect of MVPA substitution supports the interpretation that reallocating sedentary behavior to physical activity at or above a certain intensity may be beneficial to reduce the AF risk, especially among those who engaged in inadequate levels of MVPA. Meeting PA guidelines may be an effective intervention of AF risk reduction. ^27,40^ Prior studies of PA and AF also suggest that associations may vary by age and activity level and may be nonlinear.^41^ The stronger REGARDS estimates among participants aged ≥ 65 years may reflect their higher baseline AF risk or a greater relative benefit of increasing MVPA from low levels. The stronger association among participants reporting <7 hours/day of sleep may mark a less favorable cardiometabolic or 24-hour behavior profile; however, REGARDS sleep duration was self-reported, and the ≥7-hour stratum combined recommended and long sleep durations, potentially obscuring heterogeneity within that group that warrant further investigation. Reducing sedentary time alone may not be sufficient for AF prevention unless the displaced time is replaced with activity of at least moderate intensity. The results from both studies may help refine behavioral counseling by encouraging not only sitting less, but also purposeful replacement with more intense physical activity when feasible.

Key strengths of our study include objectively measured actigraphy data, which reduces self-report bias in behavior times, and the application of isotemporal substitution modeling that considers the finite 24 hours and correlations between movement times. An additional strength is the use of two population-based prospective cohorts from different countries, allowing adequate incident AF case identification and comparison across populations. A few limitations should be acknowledged. First, accelerometer protocols differed between cohorts, and neither device directly measures posture; therefore, sitting and standing may be misclassified. Sleep duration in REGARDS was self-reported because the accelerometer was removed during sleep. Self-reported sleep can differ systematically from objective sleep duration and may more closely reflect perceived sleep or time in bed, creating additional misclassification in sleep-stratified analyses.^42^ Second, the relatively low accelerometer return rate in UKB (45%) reduced the representativeness, whereas 82.1% of REGARDS ancillary study participants returned device. Third, movement behaviors were captured over a week at a single time point in both cohorts, which may be subject to change over follow-up. Fourth, residual confounding due to unmeasured factors and time-varying factors was possible, and the attenuation after adjustment may reflect confounding, overadjustment, or adjustment for factors that may lie on the causal pathway. Lastly, AF can be intermittent and asymptomatic. A study ECG may miss paroxysmal AF between episodes, while record-based ascertainment depends on healthcare contact and clinical detection; therefore, some incident AF was likely missed, and differences in ascertainment intensity may have contributed to between-cohort heterogeneity.

## CONCLUSION

In conclusion, our findings from the UKB and REGARDS suggest that replacing sedentary behavior with MVPA may decrease risk of incident AF, while shifting sedentary time to LPA and sleep does not appear to substantially influence AF risk. These results support efforts to reduce sedentary time and promote MVPA as a strategy for AF prevention among adults, particularly those with low baseline MVPA or higher baseline risk. Further investigations using repeated accelerometry assessment and additional studies to clarify the potential pathophysiology are needed.

## Data Availability

The UK Biobank will make the data available to all bona fide researchers for all types of health-related research that is in the public interest, without preferential or exclusive access for any person. All researchers will be subject to the same application process and approval criteria as specified by UK Biobank. For more details on the access procedure, see the UK Biobank website: www.ukbiobank.ac.uk.
REGARDS data are not publicly available due to ethical and legal restrictions. To abide by its obligations with NIH/NINDS and the Institutional Review Board of the University of Alabama at Birmingham, REGARDS facilitates data sharing through data use agreements. Any investigator is welcome to access the REGARDS data, including statistical code, through this process. Requests for data access may be sent to.

https://www.uab.edu/soph/regardsstudy/data-sharing

## ACKNOWLEDGEMENT

The authors thank the participants, staff, and investigators of the UK Biobank and REGARDS study for their valuable contributions. A full list of participating REGARDS investigators and institutions can be found at: https://www.uab.edu/soph/regardsstudy/.

## DISCLOSURES

EBL reports consulting fees from Pfizer and service on a data and safety monitoring board for the University of Pittsburgh.

## FUNDING

This work is supported by American Heart Association Predoctoral Fellowship (23PRE1020888, Li). The REasons for Geographic And Racial Differences in Stroke Study (REGARDS) is supported by RFI NS041588, co-funded by the National Institute of Neurological Disorders and Stroke (NINDS) and the National Institute on Aging, National Institutes of Health Department of Health and Human Service. The accelerometer study was supported by R01-NS061846 from the NINDS with additional funding provided by an unrestricted grant from the Coca-Cola Company. The content is solely the responsibility of the authors and does not necessarily represent the official views of the NINDS or the NIA.

## DATA SHARING

The UK Biobank will make the data available to all bona fide researchers for all types of health-related research that is in the public interest, without preferential or exclusive access for any person. All researchers will be subject to the same application process and approval criteria as specified by UK Biobank. For more details on the access procedure, see the UK Biobank website: www.ukbiobank.ac.uk.

REGARDS data are not publicly available due to ethical and legal restrictions. To abide by its obligations with NIH/NINDS and the Institutional Review Board of the University of Alabama at Birmingham, REGARDS facilitates data sharing through data use agreements. Any investigator is welcome to access the REGARDS data, including statistical code, through this process. Requests for data access may be sent to.

## Notes

### Competing Interest Statement

The authors have declared no competing interest.

### Author Declarations

This work is approved by IRB at Emory University (IRB2023P005425). The REGARDS study was approved by the institutional review boards of participating institutions. The data used from the UK Biobank Resource was under application number 34031.

